# Modelling the Impact of Prevention and Treatment Interventions on HIV and Hepatitis C Virus Transmission Among People Who Inject Drugs in Kenya

**DOI:** 10.1101/2021.02.02.21251008

**Authors:** Jack Stone, Hannah Fraser, Josephine G Walker, Nyashadzaishe Mafirakureva, Bernard Mundia, Charles Cleland, Bartilol Kigen, Helgar Musyoki, Wanjiru Waruiru, Allan Ragi, Parinita Bhattacharjee, Nok Chhun, John Lizcano, Matthew J Akiyama, Peter Cherutich, Ann Kurth, Niklas Luhmann, Peter Vickerman

## Abstract

**Background:** People who inject drugs (PWID) in Kenya have a high prevalence of HIV (14-26%) and HCV (11-36%). Needle and syringe programmes (NSP) and antiretroviral therapy (ART) have high coverage among PWID, while HCV treatment and opioid substitution therapy (OST) access is low.

**Methods:** A dynamic model of HIV (sexual and injecting-related) and HCV (injecting-related) transmission among PWID was calibrated using Bayesian methods to data from Nairobi and the Coastal region. We projected the impact of existing coverage levels of interventions (ART: 64-66%; OST: 4-7%; NSP: 54-56%) in each setting, and the impact over 2020-2030 of increasing the coverage of OST (50%) and NSP (75%; ‘full HR’), ART (UNAIDS 90-90-90 target), HCV treatment (1000 over 5 years), and reducing HIV sexual risk by 75%. We estimated HCV treatment levels needed to reduce HCV incidence by 90% with or without full HR.

**Findings:** Since 2013, HR has averted 15.1-20.6% (range in medians across settings) of HIV infections and 29.0-31.6% of HCV infections across Nairobi and the Coastal region, with most impact being due to NSP. Conversely, ART has only averted <5% of HIV infections since 2004 because of sub-optimal viral suppression (28-48%). Going forward, Full HR and ART could reduce HIV incidence by 58.2-62.0% and HCV incidence by 62.6-81.6% by 2030 across these settings. If sexual risk is also reduced, HIV incidence would reduce by 77.1-81.4%. Alongside full HR, treating 896 PWID over 2020-2025 could reduce HCV incidence by 90% by 2030.

**Interpretation:** Existing interventions have had moderate impact on HIV and HCV transmission in Kenya, but may have substantial impact if scaled-up. However, to achieve HIV and HCV elimination, reductions in sexual risk are needed and a scale-up in HCV treatment.

**Funding:** Global Fund, MDM

## Introduction

Kenya has a high prevalence of HIV (4.9% in 2018) among adults[1]. However, the HIV prevalence (14-20%) among people who inject drugs (PWID) far exceeds this[2, 3], particularly among females (29-61%). An estimated 7.5% of new HIV infections in Kenya are attributable to PWID, with this increasing to 18.7% on the Kenyan Coast[4].

In contrast, the prevalence of HCV among PWID in Kenya is relatively low (11-36% [5]) compared to other global settings, likely reflecting the recency of injecting drug use (IDU) in sub-Saharan Africa (SSA). Although access to diagnosis and treatment for HCV has been negligible in Kenya, recent pilot programmes among PWID have demonstrated the feasibility of such strategies in opioid substitution therapy (OST) clinics and harm reduction interventions[6]. The Kenyan government has recently secured direct acting antiviral (DAA) treatments for 1,000 people, with their national HCV guidelines recognising the importance of treating PWID.

Evidence shows that OST and needle and syringe programmes (NSP) are effective at reducing the risk of HIV[7, 8] and HCV acquisition[9], but their coverage remains low in SSA[10]. Kenya initiated NSP in 2013, with approximately half of PWID accessing NSP in 2017. OST initiated in 2014, but coverage remains low at <5%[10].

The World Health Organization (WHO) and UNAIDS have set goals for eliminating HCV and HIV by 2030. Although modelling can help guide intervention planning for reaching these goals, no such analyses have considered the IDU-related epidemics in SSA. To aid policymaking in Kenya and SSA, we used modelling to evaluate the impact of existing and scaled-up prevention and treatment interventions on the HIV and HCV epidemics among PWID in Kenya, and so considered what would be needed to reach elimination.

## Methods

### Model description

We developed a dynamic model of HIV and HCV transmission amongst PWID for two sites in Kenya: Nairobi and the Coastal region (specifically Mombasa, Kilifi and Kwale). As shown in Figure 1, we stratified the modelled population by gender, HIV infection and treatment status, HCV infection, OST and NSP status. The model equations are in the appendix.

**Figure 1:**
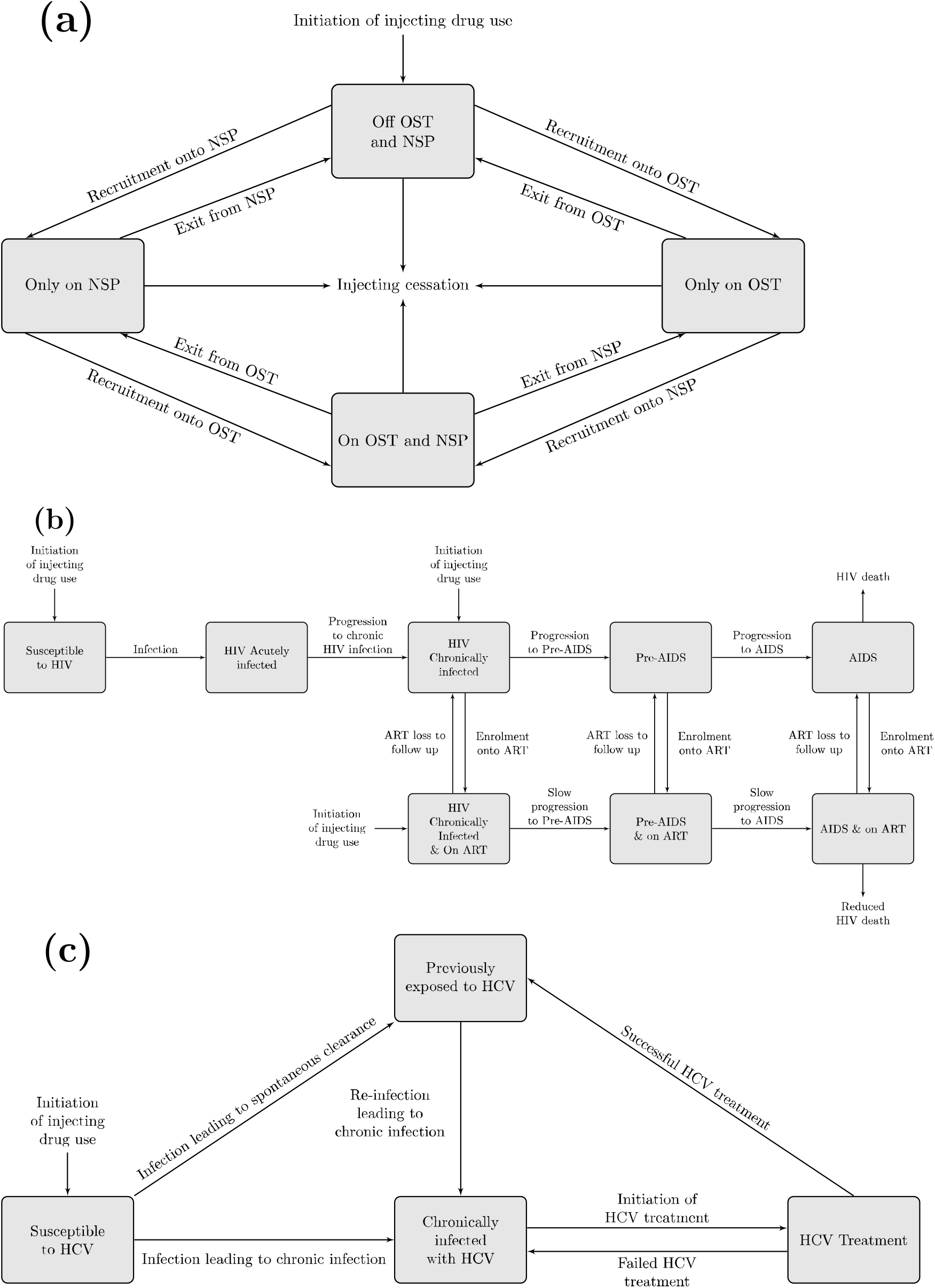
Model schematic of (a) harm reduction interventions; (b) HIV transmission and treatment; (c) HCV transmission and treatment.

Individuals enter the model through initiating IDU, susceptible to HCV, and not accessing OST or NSP. Some enter the model HIV-infected, with some on ART. Entry is balanced by individuals ceasing IDU or non-HIV/HCV related death. PWID can also experience HIV-related mortality.

Susceptible PWID become HIV and HCV infected through sharing injecting equipment, with HIV also being sexually transmitted. Injecting behaviour data and HCV prevalence patterns suggest male and female PWID have similar injecting risks (appendix); therefore, we assume that their injecting HIV and HCV transmission risk is the same. For sexual HIV transmission, we just model heterosexual transmission because few male PWID report sex with men (<2.2% last month)[2]. Differences in HIV prevalence and sexual risk behaviours (Table 1 and Appendix table 3) suggest female PWID have higher sexual HIV transmission risk than males, which we incorporate. We assume PWID have sexual partners with other PWID and the general population, with the same sexual risk behaviour assumed for each. We did not model sexual HCV transmission[11].

**Table 1:**
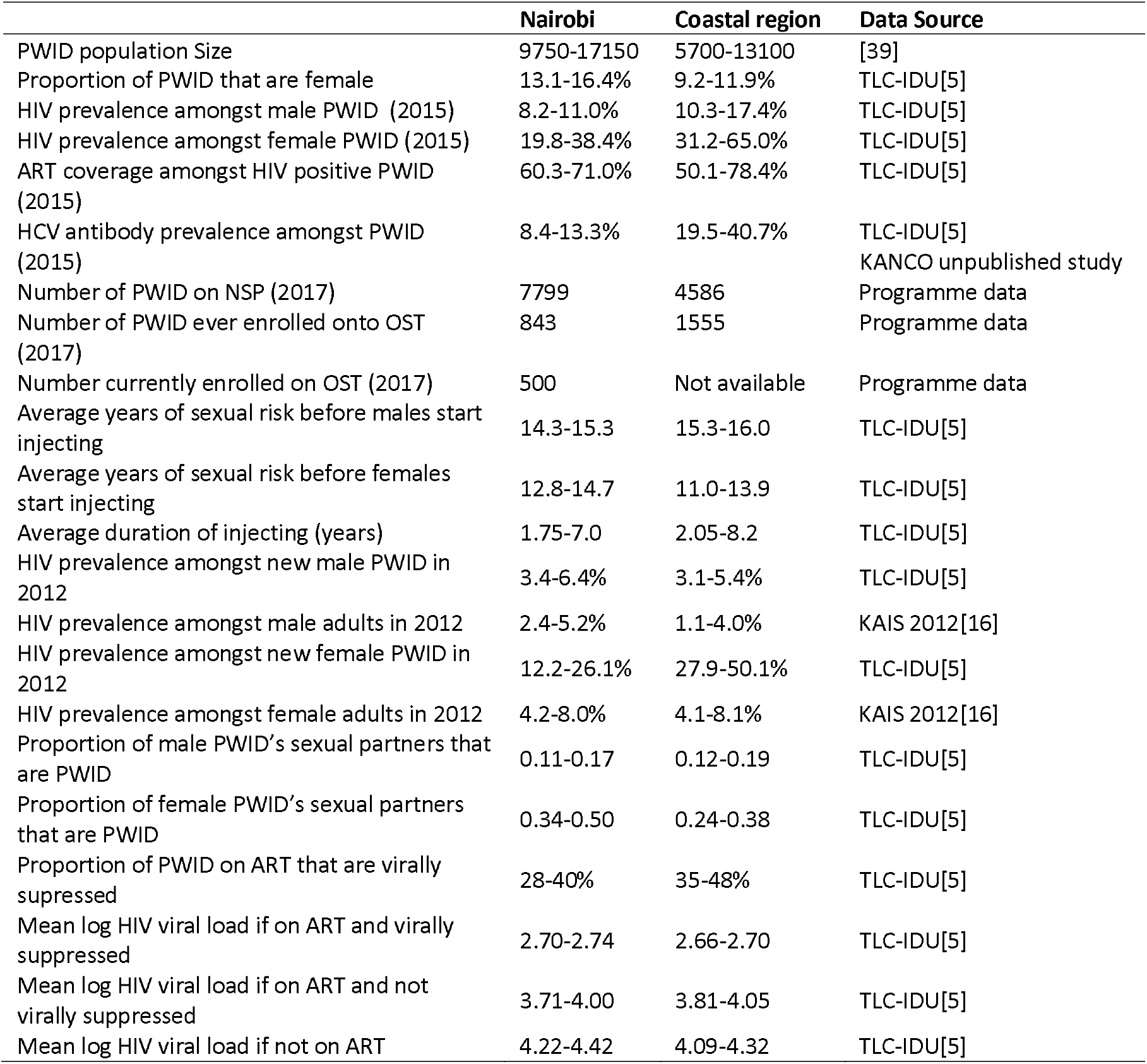
Summary of main prior parameter ranges and calibration data (most recent estimates) used for Nairobi and the Coastal region. Full parameter tables and calibration data are in the Appendix.

The risk of sexual HIV transmission is dependent upon the HIV prevalence amongst PWID and the general population, their ART coverage and the effectiveness of ART at reducing sexual HIV transmission[12]. The risk of injecting HIV transmission is dependent on the HIV prevalence among PWID, their ART coverage and its effectiveness at reducing injecting HIV transmission (assumed 0-30% less effective than for sexual transmission). The injecting HIV transmission risk is decreased for PWID on OST (on average by 54%) and/or NSP (by 58%)[7, 8]. We assume that PWID mix randomly to form potential transmission contacts with other PWID.

Following HIV infection, individuals enter a short acute phase of infection before progressing to the chronic, pre-AIDS and AIDS phases of infection at fixed rates. Individuals in the acute and pre-AIDS phases of infection have heightened infectivity[13]. Individuals in the AIDS phase experience HIV-related mortality and only engage in risk behaviours if on ART. Individuals in the chronic, pre-AIDS or AIDS phases of infection can be enrolled onto ART, which reduces disease progression and HIV infectivity. The reduction in infectivity while on ART depends on the proportion who are virally suppressed[12]. PWID receiving ART can be lost to follow-up (LTFU) and then re-enrolled at the same rate as ART-naïve PWID. Being on OST increases the likelihood of initiating ART (on average by 87%), reduces LTFU (by 23%), and improves levels of viral suppression on ART (by 45%)[14].

Similarly, the risk of HCV transmission is dependent on the chronic HCV prevalence among PWID. The HCV transmission risk is decreased for PWID on OST (on average by 50%) and/or NSP (by 56%)[9]. Following HCV infection, some PWID spontaneously clear infection (differs by HIV infection) and enter the previously exposed compartment (antibody (Ab)+, RNA-), with the remainder progressing to chronic infection (Ab+, RNA+). Chronically infected PWID can receive HCV antiviral treatment (lasts 12 weeks), with most (90.1%[6]) achieving a sustained virologic response (SVR, effective cure) and entering the previously exposed compartment. Those not achieving SVR return to the chronic HCV infection compartment, whereupon re-treatment can occur at the same rate as primary treatment. We assume that HIV-HCV co-infected PWID are more infectious than HCV mono-infected PWID[15]. No HCV-related mortality is assumed in the model due to the short duration of injecting.

### Model Parameterisation and calibration

The model was calibrated to Nairobi and the Coastal region using data from various sources, including the 2012 Kenya AIDS indicator survey (KAIS)[16], 2015 and 2016 national polling booth surveys[17], 2011 integrated bio-behavioural assessment (IBBA) among PWID in Nairobi[3], national OST and NSP programme data, and the Testing and Linkage to Care study (TLC-IDU; <u>NCT01557998</u>)[2, 5]. Most data came from the TLC-IDU study, which consisted of 6 rounds (2012-2015) of cross-sectional surveys involving respondent-driven sampling of PWID from 4 sites in Nairobi and 6 in Coastal Kenya. Participants were tested for HIV infection and HIV viral load at each round and for HCV antibody in round 6.

We assume that injecting drug use initiated between 1997-2001 in both sites[18]. OST, NSP and ART were scaled-up in line with available data (Figure 2) using different recruitment rates over specific time periods where necessary, and assuming constant coverage after the last available data estimate (see appendix). ART LTFU amongst PWID was estimated using data from the Kenyan general population[19–24], increased to account for PWID generally having greater LTFU than the general population[25].

**Figure 2:**
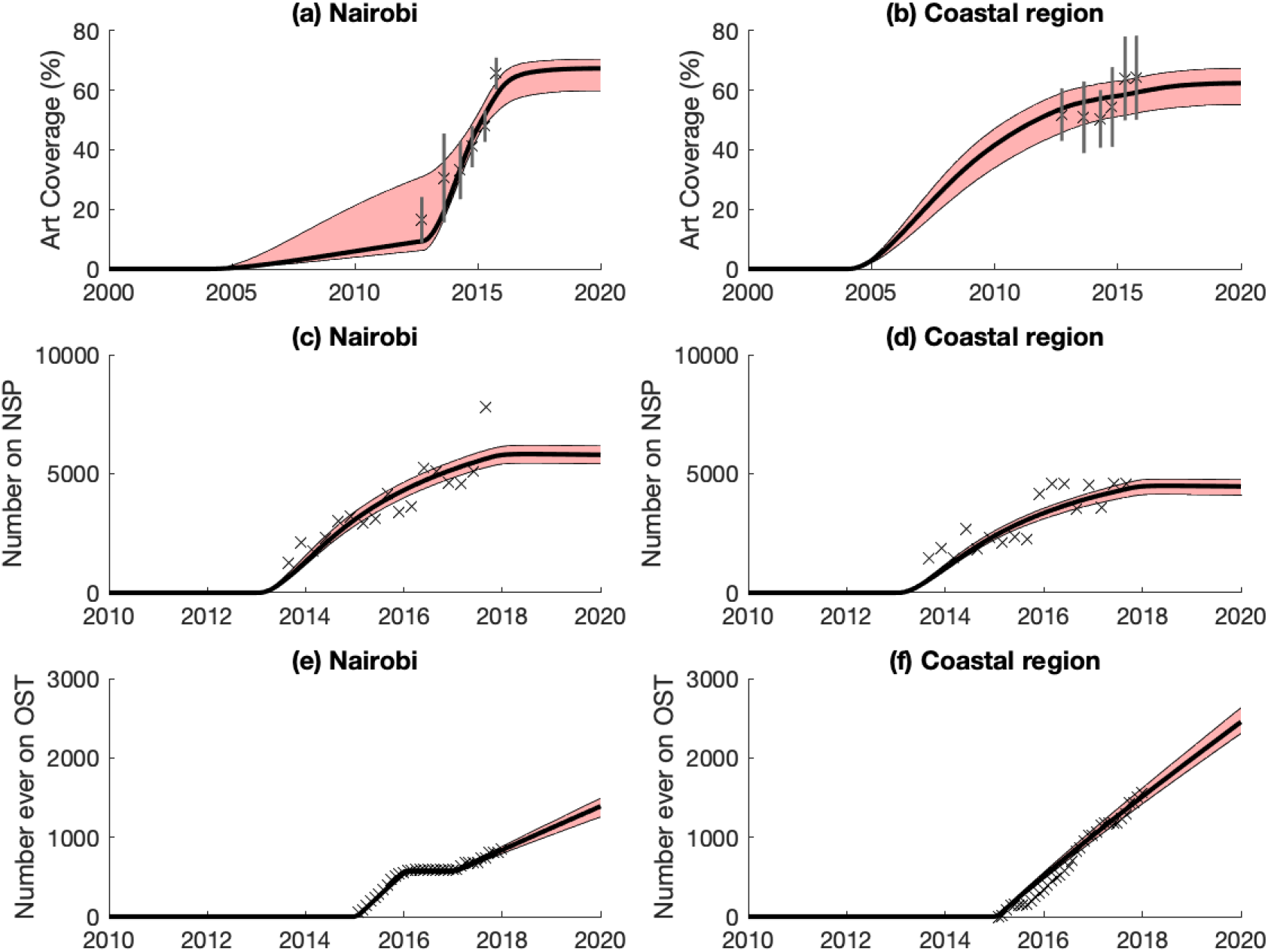
Model projections of intervention scale-up among PWID over time. (a) and (b) ART coverage in Nairobi and the Coastal region; (c) and (d) number of PWID on NSP in Nairobi and the Coastal region; (e) and (f) number of PWID ever on OST in Nairobi and the Coastal region. Black lines show the median model projections whilst the shaded area shows the 95%CrI for the baseline projections. Data points with their 95%CI (where appropriate) are shown for comparison.

For each site, the model was calibrated using an approximate Bayesian computation Sequential Monte Carlo (ABC SMC) method[26] to data on: the PWID population size, the proportion of PWID that are females, HIV prevalence amongst male and female PWID over time, HCV antibody prevalence amongst all PWID, ART coverage amongst HIV positive PWID over time, the number of PWID currently and ever enrolled onto OST over time, and the number of PWID currently in contact with NSP over time. Table 1 summarises prior ranges for key parameters and calibration data, with full details in the appendix.

Using all 6 rounds of the TLC-IDU study, we used logistic regression to estimate the HIV prevalence in 2013 among male PWID when they initiate injecting. This was sampled in the ABC SMC and estimated over time by assuming the same ratio between the HIV prevalence among new male PWID and general population males over time. The sampled HIV prevalence among new male PWID in 2013 was also used to estimate a range for the average level of sexual HIV transmission risk experienced by males over the duration of sexual risk before they start injecting (using TLC-IDU data). The same level of sexual HIV transmission risk was assumed after they initiate injecting, with this range being calibrated to in the ABC. The same method was used for female PWID.

We performed multiple iterations of the ABC SMC method, each consisting of 5,000 parameter sets, until the main results from the combined parameter sets converged (appendix). These combined parameter sets were sampled, weighted by their likelihood (appendix), to give a final group of baseline model runs/fits consisting of 10% of the combined parameter sets (3,000 for Nairobi, 4,000 for Coast); these were used in subsequent analysis.

## Model analyses

### Impact of existing interventions

We projected the baseline HIV and HCV epidemics among PWID until 2019, including existing interventions, and estimated the percentage decrease in HIV infections in 2019 if there was no sexual HIV transmission. We then projected the proportion of infections prevented by each intervention from the date that they were first implemented to 2019; 2004 for ART, 2013 for NSP and 2015 for OST. This was done by comparing the number of HIV and HCV infections over these periods with a counterfactual where the efficacy of specific interventions is set to zero.

### Impact of scaling-up interventions

We projected the baseline HIV and HCV epidemics among PWID until 2030, assuming no future scale-up of interventions. We then projected the impact, in terms of reduction in HIV/HCV incidence and prevalence and proportion of new infections averted (over 2019-2030), of scaling-up harm reduction interventions and/or HIV and HCV treatment from 2019. Specifically, we modelled the following scenarios:

1. **Full harm reduction (HR)**: Scale-up OST and NSP to 50% and 75% coverage, respectively;
2. **Full ART**: Scale-up ART to meet the UNAIDS 90-90-90 targets among PWID, i.e. 81% ART coverage amongst HIV-infected PWID with 90% virally supressed;
3. **Full HR+Full ART**: Full HR and Full ART;
4. **Treat HCV**: Treat 1,000 HCV-positive PWID over next 5 years (200 per year);
5. **Full HR+Treat HCV**: Full HR plus scaling-up HCV treatment as in (4).

We also estimated the additional impact on HIV of reducing HIV sexual transmission risk by 75% from 2019 and the required HCV treatments for achieving the WHO HCV elimination target of decreasing HCV incidence by 90% over 2019-2030.

### Uncertainty analysis

To determine which parameter uncertainties are important for contributing to the variability in our model projections, a linear regression analysis of covariance was performed on the relative reduction in HIV and HCV incidence achieved over 2019-2030 from Full HR+Full ART. The proportion of the model outcome’s sum-of-squares contributed by each parameter was calculated to estimate the importance of individual parameters to the overall uncertainty.

## Results

### Baseline Model Projections

The calibrated model fit the data well (Figures 2 and 3), suggesting a slowly decreasing HIV epidemic among PWID in both settings, an increasing HCV epidemic in Nairobi and slowly decreasing HCV epidemic in Coastal region. In both settings, up to 5-times higher HIV prevalences are projected among female PWID than male PWID.

**Figure 3:**
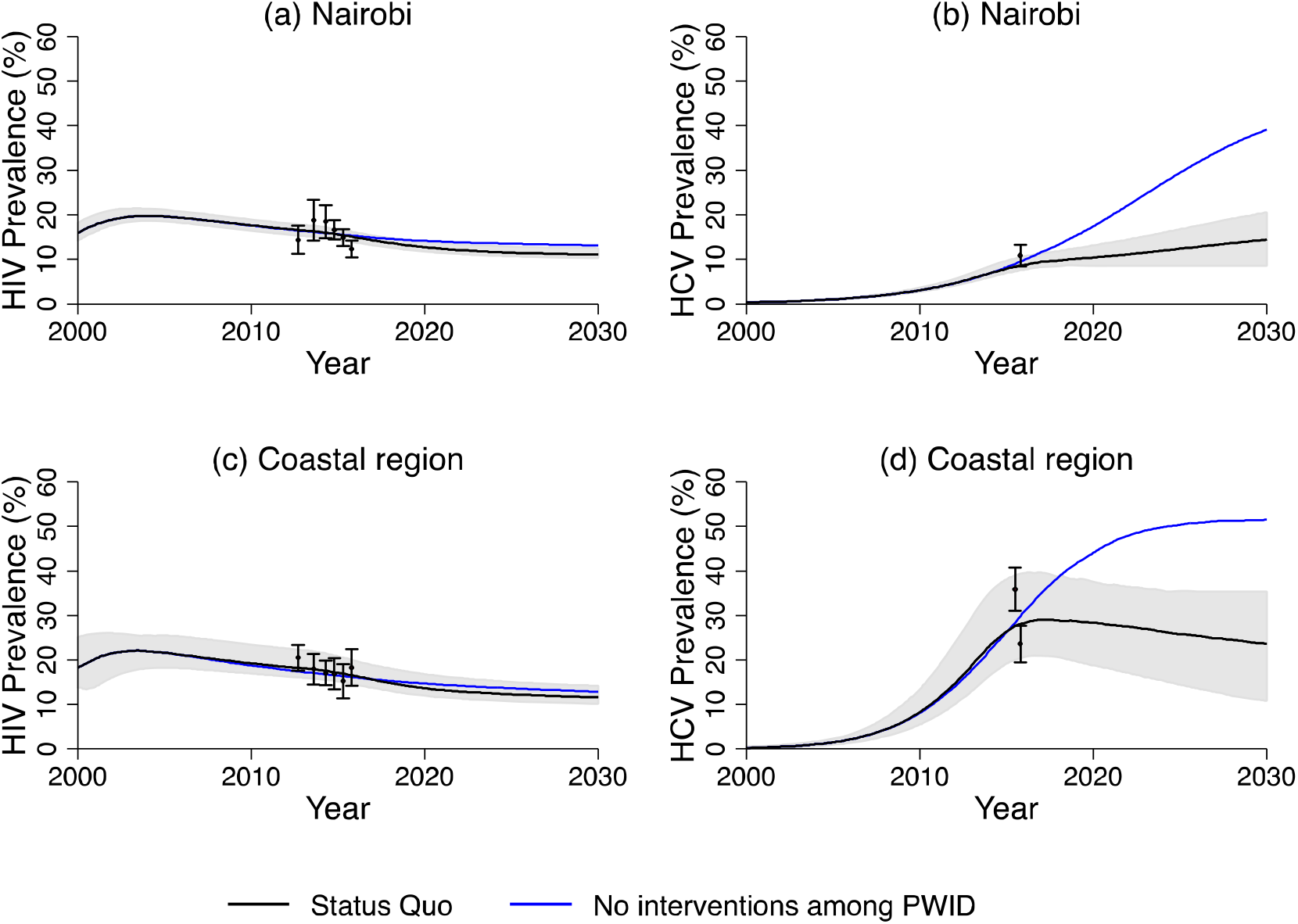
Model projections of HIV and HCV prevalence among PWID over time with and without the impact of existing interventions. (a) HIV prevalence in Nairobi; (b) HCV antibody prevalence in Nairobi; (c) HIV prevalence in the Coastal region; and (d) HCV antibody prevalence in the Coastal region. All lines show the median model projections whilst the shaded area shows the 95%CrI for the status quo projections. Data points with their 95%CI are shown for comparison.

The model projects HIV and HCV incidences of 2.1 (95%CrI: 1.6-2.5) and 3.6 (95%CrI: 2.4-4.8) per 100py in 2019 in Nairobi, respectively, and 2.3 (95%CrI: 1.8-3.5) and 12.2 (95%CrI: 7.3-17.4) per 100py in Coastal region. In both settings, sexual HIV transmission is an important mode of HIV transmission, with 31.0% (95%CrI: 26.0-55.4) and 65.4% (95%CrI: 61.0-84.3) of new HIV infections among male and female PWID being due to sexual transmission in Nairobi, respectively, compared to 16.8% (95%CrI: 9.4-25.8) and 68.0% (95%CrI: 54.7-76.5) in Coastal region. Overall, the HIV incidence among female PWID is over double that of male PWID in both Nairobi (IRR: 2.0, 95%CrI: 1.9-2.9) and Coastal region (IRR: 2.6, 95%CrI: 1.9-3.2), with the incidence of sexually acquired HIV infection being 4-times higher among female PWID than male PWID in Nairobi (IRR: 4.34, 95%CrI: 3.8-4.9) and 10-times higher in Coastal Region (IRR: 10.6, 95%CrI; 8.3-13.1).

### Impact of existing interventions

In 2019, for Nairobi and Coastal region, the model projects NSP coverages of 53.5% (95%CrI: 50.2-63.6) and 55.5% (95%CrI: 47.4-59.9), OST coverages of 3.7% (95%CrI: 2.5-4.2) and 7.1% (95%CrI: 5.9-8.7), and ART coverages of 67.3% (95%CrI: 59.8-70.4) and 62.3% (95%CrI: 55.1-67.2), respectively. Since their implementation, these interventions have had similar impact in each setting, with more impact achieved on HCV (Figure 4). For instance, harm reduction interventions (since 2013) have averted 15.1% (95%CrI: 9.7-21.2) and 29.0% (95%CrI: 25.3-44.0) of new HIV and HCV infections in Nairobi, while 20.6% (95%CrI: 13.3-26.3) and 31.6% (95%CrI: 23.2-42.2) were averted in Coastal region. The impact of NSP and OST on HIV is less than for HCV because of the high levels of sexual HIV transmission among PWID, while NSP had more (6-10-times) impact than OST(Figure 4) because of its much higher coverage.

**Figure 4:**
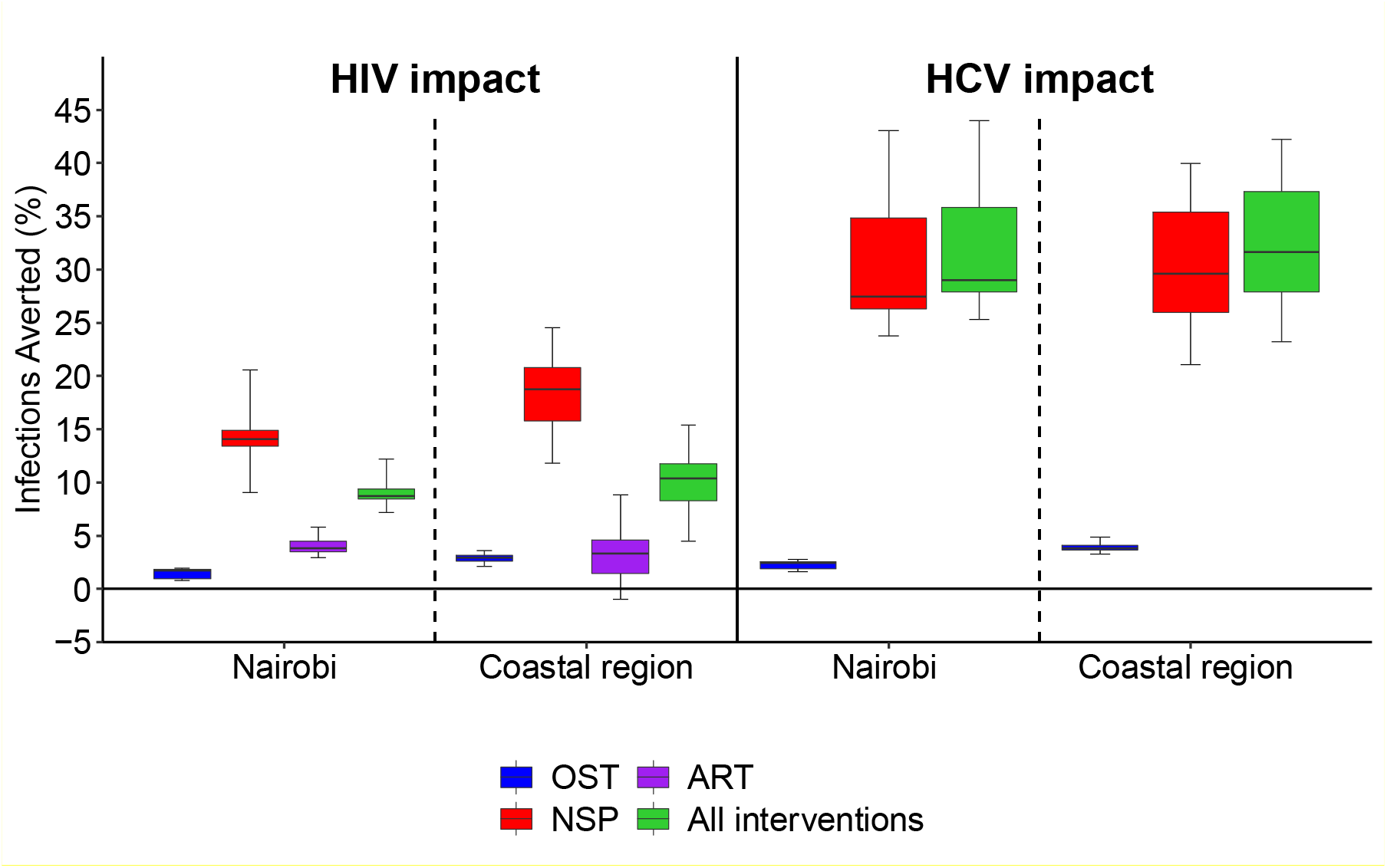
Proportion of new HIV and HCV infections averted among PWID in Nairobi and the Coastal region by existing interventions from the start of their implementation to 2019. Boxes indicate the interquartile range, with the lines inside indicating the median impact, with whiskers representing 95% CrI for the simulations.

In contrast, ART (among PWID) has averted much fewer HIV infections since its introduction in 2004, averting 3.8% (95%CrI: 3.0-5.8) and 3.4% (95%CrI: −1.0-8.9) of HIV infections in Nairobi and the Coastal region, respectively. This low impact is primarily due to the low levels of viral suppression in both settings (28-40% in Nairobi and 35-48% in Coastal region) resulting in the efficacy of ART for reducing infectivity being low (posteriors 39-55% for sexual HIV transmission and 36-41% for injecting), paired with increased survival while on ART (by ∼4-times) and the slow scale-up of ART in the early years (Figure 2).

Figure 3 shows that without these interventions, HCV prevalence would have been much higher in both settings, while the effects on HIV prevalence are much less pronounced, partly because ART reduces HIV mortality and also because the smaller impact of the interventions on HIV incidence. HIV prevalence still declines without these interventions due to ongoing reductions in HIV prevalence among the general population.

### Impact of scaling-up interventions

Scaling-up harm reduction and ART could reduce HIV incidence by 58.2% (95%CrI: 46.4-61.6) in Nairobi and 62.0% (95%CrI: 54.6-69.6) in Coastal region over 2019-2030 (Figure 5). Just scaling-up ART or harm reduction has less impact; in Nairobi, for example, HIV incidence reduces by 45.2% (95%CrI: 28.1-49.3) through just scaling-up of harm reduction or 36.7% (95%CrI: 31.1-39.6) through just scaling-up ART. To decrease HIV incidence further in both settings, reductions in sexual risk are also required (Figure 5). Reducing sexual risk by 75% alongside scaling-up harm reduction and ART reduces HIV incidence by 81.4% (95%CrI: 79.5-88.4) in Nairobi and 77.1% (95%CrI: 73.1-82.7) in Coastal region.

**Figure 5:**
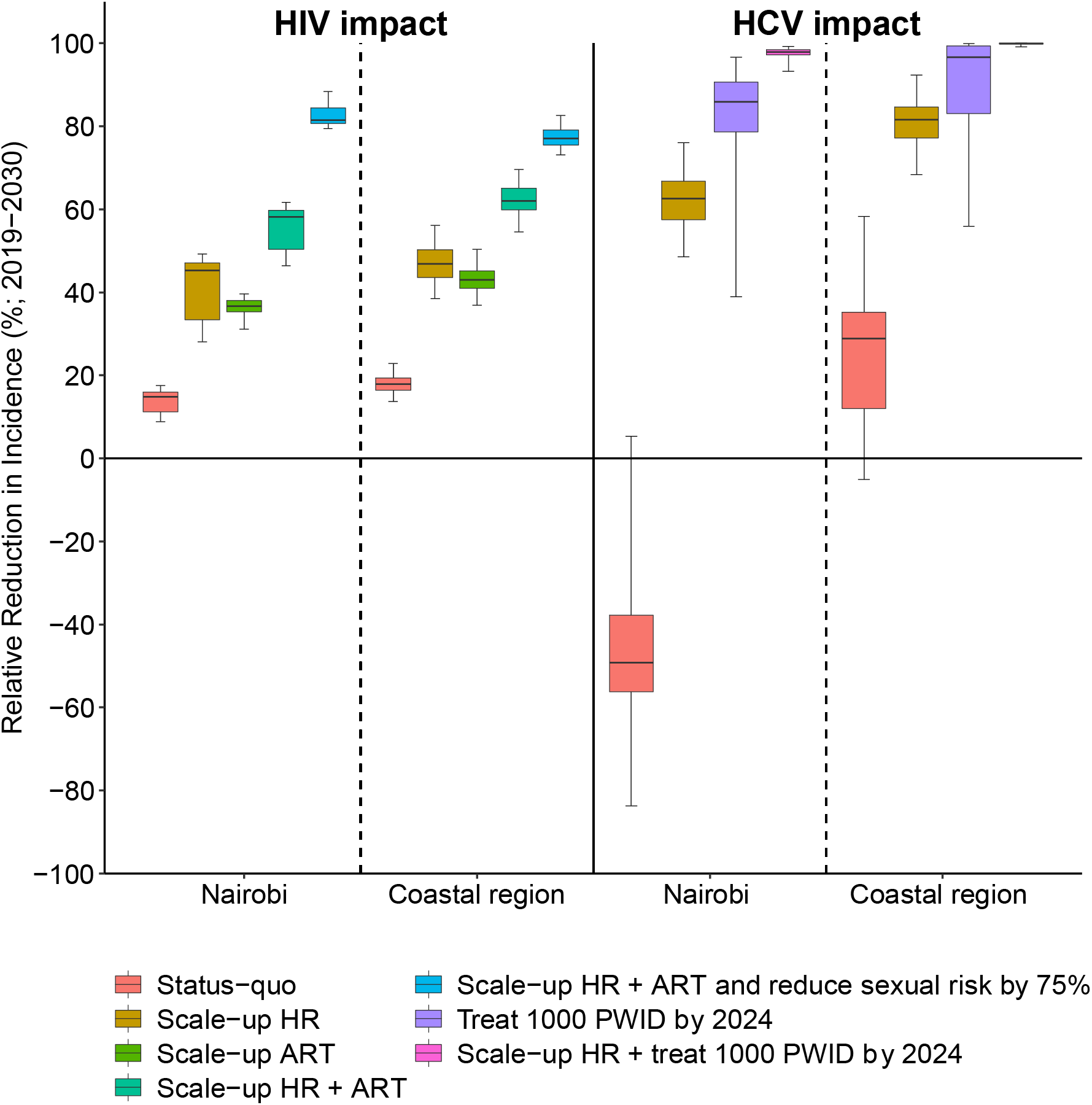
Relative reduction in HIV and HCV incidence over 2019-2030 among PWID in Nairobi and the Coastal region for different intervention scale-up scenarios. Boxes indicate the interquartile range, with the lines inside indicating the median impact, with whiskers representing 95% CrI for the simulations.

Without any scale-up of interventions, HCV incidence is projected to increase by 49.2% (95%CrI: −5.3-83.8) in Nairobi over 2019-2030, and decrease by 28.8% (95%CrI: −5.1-58.3) in Coastal region; both with considerable uncertainty. In both settings, scaling-up OST and NSP could reduce HCV incidence by 62.5% (95%CrI: 48.5-76.0) in Nairobi and 81.6% (95%CrI: 68.4-92.4) in Coastal region (Figure 5). Conversely, treating 1,000 PWID in each setting over the next 5 years could reduce HCV incidence by 85.9% (95%CrI: 39.0-96.6) in Nairobi and 96.7% (95%CrI: 55.9-99.9) in Coastal Region, while combining this with scaled-up OST and NSP could nearly eliminate HCV. A 90% reduction in HCV incidence is achieved by 2030 if alongside scaling-up harm reduction, 622.1 (95%CrI: 359.3-909.6) PWID are treated by 2024 in Nairobi and 273.8 (95%CrI: 0.0-621.0) are treated in Coastal region, with this increasing to 1,106.2 (679.8-1,784.4) and 872.1 (361.7-1,440.9) with no scale-up of OST and NSP.

### Uncertainty analysis

Analyses of covariance indicate that uncertainty in the levels of sexual transmission (15.3% and 36.6% of uncertainty in Nairobi and Coastal region, respectively), effectiveness of NSP (Nairobi 51.5%, Coastal region: 20.7%) and baseline (2019) levels of HIV incidence (Nairobi: 17.3%; Coastal region: 18.9%) contributed most to the variability in the impact of scaling-up harm reduction interventions and ART on HIV incidence. Conversely, uncertainty in the effectiveness of NSP (Nairobi: 50.4%, Coastal region: 46.6%), duration of injecting (Nairobi: 22.0%, Coastal region: 47.7%) and NSP coverage (Nairobi: 12.7%, Coastal region: 3.1%) contributed most to the variability in the impact of scaling-up harm reduction on HCV incidence.

## Discussion

### Main Findings

In Kenya, increases in injecting drug use since the 1990s have resulted in high HIV prevalence and expanding HCV epidemics among PWID. To inform policymaking in Kenya and SSA, we evaluated the impact of existing and future interventions for PWID in Kenya. Our projections show that only NSP so far has resulted in substantial impact, due to its rapid expansion to 54-56% coverage, preventing 14-19% of HIV infections and 27-30% of HCV infections since 2011. In contrast, the impact of ART and OST has been small among PWID (<5% of infections averted) because of sub-optimal viral suppression on ART and low coverage levels of OST. To increase impact further, harm reduction interventions and ART need to be scaled-up, with HIV incidence decreasing by about 60% and HCV incidence by 63-82% if NSP is increased to 75% coverage, OST to 50% coverage and the 90/90/90 targets for ART are reached. Beyond this, HIV incidence can only be reduced further through implementing interventions that reduce sexual risk, while HCV incidence can be reduced by 90% or more through scaling-up DAAs alongside scaling-up harm reduction interventions.

### Strengths and Limitations

The strengths of our analyses include the use of detailed epidemiological, behavioural and programmatic data from multiple sources to calibrate our model within a Bayesian framework; increasing the rigour of our analyses. We also modelled two settings within Kenya, so increasing the robustness and generalisability of our findings. However, whilst we had detailed data on trends in HIV prevalence among PWID for both settings, HCV prevalence data were limited, as for much of SSA[27]. Better data on HCV prevalence trends among PWID will improve the accuracy of our impact projections for different interventions. We used global systematic reviews to obtain estimates for the effectiveness of OST and NSP in reducing HIV and HCV transmission risk[9]. However, these reviews are largely based on studies from Europe, Australia and North America and so it is uncertain whether these interventions would have similar effectiveness in SSA. As harm reduction interventions are scaled-up, it is important to investigate whether these interventions have a similar effectiveness in SSA. When modelling the sexual transmission of HIV among PWID, we were limited by having insufficient data on numbers of sexual partners and frequency of sexual acts. Because of this we had to model the level of sexual risk indirectly. Despite these limitations, the calibrated models accurately captured the observed differences in HIV prevalence trends between male and female PWID, which is largely thought to be due to differences in sexual risk.

### Comparisons with existing studies

To our knowledge, this study, along with an accompanying study in Dar es Salaam[28], Tanzania, represents the first dynamic modelling of ongoing HIV and HCV epidemics among PWID in SSA. Previous modelling for Nairobi suggested that scaling-up of OST to 40% coverage in Kenya could reduce HIV incidence by a fifth over 10 years[29]. Adding to this, we show that OST has had limited impact so far because coverage remains low, but could have large impact if scaled-up. Other much simpler modelling studies for Kenya[30], as well as modelling from other settings[31], have shown that a combined approach of scaling-up harm reduction alongside ART is needed to substantially reduce HIV transmission. Although our modelling agrees with this, through capturing sexual HIV transmission among PWID, we also show the importance of implementing interventions to reduce sexual risk behaviours among PWID for achieving substantial reductions in HIV incidence in Kenya. This is likely to be relevant to other SSA settings. As for Dar es Salaam[28, 32] and other more established HCV epidemics[33], our modelling also suggests that scaling-up harm reduction interventions alone cannot achieve HCV elimination, but that HCV treatment is also needed.

## Conclusions

Kenya has seen an increase in injecting drug use, with harm reduction interventions only recently being implemented. Although our findings suggest the rapid scale-up of NSP has impacted on HIV and HCV transmission among PWID in Kenya, low coverage levels of OST and sub-optimal viral suppression for ART has severely limited the impact of these interventions. Very few PWID have also received DAA HCV treatment. Although OST, HCV treatment and ART to a lesser extent urgently need scaling-up among PWID in Kenya to improve impact, specific interventions are also required to improve and ensure their treatment outcomes. Also, to achieve the modelled scale-up of HCV treatment, HCV testing needs expanding, with only 22% of PWID reporting being tested for HCV in last 3 months compared to 74% for HIV.[17] This testing could occur through existing harm reduction interventions or OST clinics as demonstrated by two recent pilot HCV screening and treatment interventions in Kenya, which both achieved a high proportion (82-95%) of diagnosed PWID completing treatment[6]. Expanding OST would also have other benefits through reducing drug-related mortality as well as improving HIV and HCV testing and treatment outcomes among PWID[14, 34, 35]. This could also improve the effectiveness of HIV and HCV treatment for reducing HIV and HCV transmission[36].

For reaching WHO’s HCV elimination targets, our modelling suggests that 1,978 HCV treatments are needed among PWID by 2024 if there is no harm reduction scale-up, and 896 if harm reduction interventions are scaled-up. Conversely, the UNAIDS HIV elimination targets can only be achieved with additional interventions to dramatically reduce levels of sexual risk among PWID, particularly among female PWID. This should firstly focus on intensive promotion of condom use, which is very low (13-14% at last sex act) among main partners and only moderate (46-57%) among casual partners[2]. HIV pre-exposure prophylaxis (PrEP) could also be introduced, which could be particularly important for the large portion (39%) of female PWID engaged in transactional/commercial sex[17] but not engaged in existing interventions for female sex workers. Although PrEP is currently recommended as a prevention choice for PWID in Kenya[37], the knowledge of PrEP among female PWID will need improving[38] to ensure PrEP can have impact. Interventions should also consider non-injecting drug users, as newly initiating injectors already have high HIV prevalence, suggesting considerable sexual HIV transmission occurs before initiating injecting drug use. This may be linked to the long duration (10-20 years) of non-injecting drug use[3] prior to initiating injecting, suggesting that harm reduction interventions should focus on all drug users and reducing both their injecting and sexual transmission risk.

## Data Availability

Model code will be shared with researchers who provide a methodologically sound proposal approved by Dr Jack Stone and Professor Peter Vickerman. Proposals should be directed to; requesters will need to sign a data access agreement.

## Authors contributions

PV undertook the initial conceptualization with NL, which was refined with EW. JS developed the model with input from JGW, HF and PV. JS performed the model analyses and wrote the first draft of the manuscript. PV supervised the project. PB, HM, AK, and JGW provided data and/or undertook data analyses for the model. All authors contributed to data interpretation, writing the manuscript and approved the final version

## Acknowledgements

This study was funded by the Global Fund East Africa Harm Reduction Project and Medicins du Monde. JS, PV, and HF acknowledge support from the NIHR Health Protection Research Unit in Behavioural Science and Evaluation and from NIDA (R01AI147490). MJA acknowledges support from NIDA (R00DA043011). Funding for study data provided from AK and PC was from NIDA 5R01DA032080.

