## Appendix for "Modelling the Impact of Prevention and Treatment Interventions on HIV and Hepatitis C Virus Transmission Among People Who Inject Drugs in Kenya"

### Model equations

Let $X_{i,j,k}^{m,n}$ be the number of PWID in the model where

- subscript $i$ denotes gender ($i=1$, male; $i=2$, female)
- subscript $j$ denotes harm reduction intervention status ($j=1$, not accessing OST or NSP, never been on OST; $j=2$, accessing OST only; $j=3$, accessing NSP only, never been on OST; $j=4$, accessing both OST and NSP; $j=5$, not accessing OST or NSP, ever been on OST; $j=6$, accessing NSP only, ever been on OST)
- subscript $k$ denotes ART treatment ($k=1$, not on ART; $k=2$, on ART)
- superscript $m$ denotes HIV infection status ($m=1$, susceptible to HIV; $m=2$, acute HIV infection; $m=3$, chronic HIV infection, $m=4,$ pre-AIDS phase of infection; $m=5$, AIDS phase of infection
- superscript $n$ denotes HCV infection status ($n=1$, susceptible to HCV; $n=2$, exposed/previously infected with HCV (Ab+, RNA -ve); $n=3$, chronic HCV infection (Ab+, RNA +ve); $n=4$, HCV treatment)

The ordinary differential equation models can be written as

$$\frac{dX_{i,j,k}^{m,n}}{dt}=\Theta_{i,j,k}^{m,n}+\Sigma_{i,j,k}^{m,n}+\Lambda_{i,j,k}^{m,n}+P_{i,j,k}^{m,n}+K_{i,j,k}^{m,n}+{\Pi_{i,j,k}^{m,n}+M}_{i,j,k}^{m,n}$$

where

- $\Theta_{i,j,k}^{m,n}$represents recruitment of PWID into the model
- $\Sigma_{i,j,k}^{m,n}$ represents transitions between harm reduction states (OST and NSP)
- $\Lambda_{i,j,k}^{m,n}$ represents HIV transmission
- $P_{i,j,k}^{m,n}$ represents HIV disease progression
- $K_{i,j,k}^{m,n}$ represents transitions on and off ART
- $\Pi_{i,j,k}^{m,n}$ represents HCV transmission and treatment
- $M_{i,j,k}^{m,n}$ represents non-HIV related mortality from the model and injecting cessation

Each of these terms are described in more detail below.

##### Inflow of injectors - $\Theta_{i,j,k}^{m,n}$

The inflow of new injectors is given by

$$\Theta_{i,1,1}^{1,1}=\left( 1-p_{i} \right)\theta_{i}\left( t \right)$$

$$\Theta_{i,1,1}^{3,1}=p_{i}\left( 1-q \right)\theta_{i}(t)$$

$$\theta_{i,1,2}^{3,1}=p_{i}q\theta_{i}(t)$$

$$\Theta_{i,j,k}^{m,n}=0, for all other m,n,i,j,k$$

where

- $p_{i}$ is the HIV prevalence among PWID of gender $i$ when they initiate injecting
- $\theta_{i}$ is the number of new PWID of gender $i$ who enter the model at time $t$ – note this is set to equal the number who leave the model due to non-HIV related mortality.
- $q$ is the proportion of HIV positive new PWID who are on ART.

##### Transitions between harm reduction states - $\Sigma_{i,j,k}^{m,n}$

Transitions between harm reduction states are given by:

$$\Sigma_{i,1,k}^{m,n}=-\left( \kappa+\beta\right)X_{i,1,k}^{m,n}+\epsilon X_{i,3,k}^{m,n}$$

$$\Sigma_{i,2,k}^{m,n}=-\left( \gamma+\beta\right)X_{i,2,k}^{m,n}+\kappa(X_{i,1,k}^{m,n}+X_{i,5,k}^{m,n})+\epsilon X_{i,4,k}^{m,n}$$

$$\Sigma_{i,3,k}^{m,n}=-\left( \epsilon+\kappa\right)X_{i,3,k}^{m,n}+\beta X_{i,1,k}^{m,n}$$

$$\Sigma_{i,4,k}^{m,n}=-\left( \epsilon+\gamma\right)X_{i,4,k}^{m,n}+\kappa(X_{i,3,k}^{m,n}+X_{i,6,k}^{m,n})+\beta X_{i,2,k}^{m,n}$$

$$\Sigma_{i,5,k}^{m,n}=-\left( \kappa+\beta\right)X_{i,5,k}^{m,n}+\gamma X_{i,2,k}^{m,n}+\epsilon X_{i,6,k}^{m,n}$$

$$\Sigma_{i,6,k}^{m,n}=-\left( \epsilon+\kappa\right)X_{i,6,k}^{m,n}+\gamma X_{i,4,k}^{m,n}+\beta X_{i,5,k}^{m,n}$$

where

- $\kappa$ denotes the rate of recruitment onto OST.
- $\beta$ denotes the rate of recruitment onto NSP.
- $\gamma$ denotes the OST leaving rate.
- $\epsilon$ denotes the NSP leaving rate.

##### HIV transmission - $\Lambda_{i,j,k}^{m,n}$

The terms in this expression are concerned with HIV transmission and are given by

$$\Lambda_{i,j,1}^{1,n}=-\left( \Lambda_{i,j}^{sex}+\Lambda_{j}^{inj} \right)X_{i,j,1}^{1,n}$$

$$\Lambda_{i,j,1}^{2,n}=\left( \Lambda_{i,j}^{sex}+\Lambda_{j}^{inj} \right)X_{i,j,1}^{1,n}$$

$\Lambda_{i,j,k}^{m,n}=0$ if $m\geq3$; $\Lambda_{i,j,2}^{1,n}=0; \Lambda_{i,j,2}^{2,n}=0$

where

- $\Lambda_{i,j}^{sex}$ is the HIV sexual force of infection for PWID of gender $i$ in harm reduction state $j$ (see section below).
- $\Lambda_{j}^{inj}$ is the HIV injecting force of infection for PWID in harm reduction state $j$ (see section below).

**HIV injecting force of infection**

The HIV injecting force of infection for PWID in each intervention state $j$ is denoted by $\Lambda_{j}^{inj}$ and is given by

$$\Lambda_{j}^{inj}={\phi_{j}^{HIV}\beta}_{inj}^{HIV}Y$$

where

$$Y=\frac{\sum_{i} \sum_{n} \sum_{j} Y_{j}}{\sum_{i} \sum_{n} \sum_{j} Z_{j}}$$

and

$$Y_{j}={\Phi_{j}^{HIV}[\phi_{A}X}_{i,j,1}^{2,n}+X_{i,j,1}^{3,n}+\phi_{P}X_{i,j,1}^{4,n}+\delta_{j}^{inj}(X_{i,j,2}^{3,n}+\phi_{P}{(X}_{i,j,2}^{4,n}+X_{i,j,2}^{5,n}))]$$

$$Z_{j}=\Phi_{j}^{HIV}\left[ \sum_{m=1:4} X_{i,j,1}^{m,n}+\sum_{m=2:5} X_{i,j,2}^{m,n} \right]$$

where

- $\beta_{inj}^{HIV}$ denotes the HIV transmission rate for injecting for PWID who are not accessing OST or NSP and are not currently on ART.
- $\Phi_{j}^{HIV}$ denotes the relative reduction in HIV transmission for injecting transmission if accessing OST ($j=2$), NSP $(j=3,6)$ or both $\left( j=4 \right)$. Note that when not accessing OST or NSP $(j=1,5)$ $\Phi_{j}^{HIV}=1$.
- $\phi_{A}$ denotes the relative increase in HIV transmissibility if in the acute stage of infection compared to the latent stage of infection.
- $\phi_{P}$ denotes the relative increase in HIV transmissibility if in the pre-AIDS stage of infection (or AIDS stage when on ART) compared to the latent stage of infection.
- $\delta_{j}^{inj}$ denotes the average reduction in HIV transmission through injecting by ART for PWID off OST $(j=1,3,5,6)$ or on OST ($j=1,2$). Note that $\delta_{j}$ is explained further below.

**HIV sexual force of infection**

The HIV sexual force of infection for PWID of gender $i$ is denoted by $\Lambda_{i}^{sex}$ and is given by

$$\lambda_{1,j}^{sex}={\beta_{2}^{sex}(q}_{1}Q_{2}+\left( 1-q_{1} \right)r_{2}(1-g+g\delta_{2}^{sex})$$

$$\lambda_{2,j}^{sex}=\beta_{1}^{sex}(q_{2}Q_{1}+\left( 1-q_{2} \right)r_{1}(1-g+g\delta_{2}^{sex})$$

where

$$Q_{i}=\frac{\sum_{n} \sum_{j} L_{i,j}^{N}}{\sum_{n} \sum_{j} L_{i,j}^{D}}$$

and

$$L_{i,j}^{N}={\phi_{A}X}_{i,j,1}^{2,n}+X_{i,j,1}^{3,n}+\phi_{P}X_{i,j,1}^{4,n}+\delta_{j}^{inj}(X_{i,j,2}^{3,n}+\phi_{P}{(X}_{i,j,2}^{4,n}+X_{i,j,2}^{5,n}))$$

$$L_{i,j}^{D}=\sum_{m=1:4} X_{i,j,1}^{m,n}+\sum_{m=2:5} X_{i,j,2}^{m,n}$$

where

- $\beta_{i}^{sex}$ denotes the sexual HIV transmission rate from group $i$ to the opposite gender for PWID who are not currently on ART.
- $q_{i}$ is the proportion of sexual partners of group $i$ that are with other PWID.
- $r_{i}$ is the HIV prevalence among general population of gender $i$
- $g$ is the ART coverage in the general population
- $\delta_{j}^{sex}$ denotes the average reduction in HIV transmission through sexual risk by ART for PWID off OST $(j=1,3,5,6)$ or on OST ($j=2,4$). Note that $\delta_{j}$ is explained further below.

Effectiveness of ART for reducing HIV sexual and injecting transmission

If $V_{j}^{S}=p_{s} (j=1,3,5,6)$ denotes the proportion of PWID on ART but not OST who are virally supressed, and $r_{s}$ denotes the odds ratio of viral suppression when on OST compared to PWID not on OST then the proportion of PWID on ART with viral suppression when on OAT is given by

${V_{j}^{S}=p}_{s}^{*}=\frac{p_{s}r_{s}}{1+p_{s}\left( r_{s}-1 \right)},$ $j=2,4$.

To determine the decrease in HIV transmission risk among virally suppressed and unsuppressed PWID on ART, we estimated the log difference between the baseline plasma viral load ($PVL-v_{b})$ for PWID off ART, and PWID on ART with suppressed ($v_{s})$ PVL $\Delta_{s}=v_{b}-v_{s}$, or unsuppressed ($v_{u})$ PVL$\Delta_{u}={v_{b}-v}_{u}$. Because prior studies [1, 2] suggest HIV transmission risk increases (factor $r_{t}$) for each log increase in PVL, these log differences in PVL were used to estimate the relative decrease in transmission risk among virally suppressed ${(e}_{s})$ and unsuppressed ${(e}_{u})$ PWID on ART:

$$\begin{aligned} e_{s}=1/{(r}_{t}^{\Delta_{s}}) \\ e_{u}=1/{(r}_{t}^{\Delta_{u}}) \end{aligned}$$

The average reduction in HIV sexual transmission by ART for PWID off OST $(j=1,3,5,6)$ or on OST $(j=2,4)$ is then given by

$${\delta_{j}^{sex}=V}_{j}^{S}e_{s}+\left( 1-V_{j}^{S} \right)e_{u}$$

If $F_{inj}$ is the relative effectiveness on viral suppression of ART for injecting compared to sexual transmission then the average reduction in HIV injecting transmission for PWID off $(j=1,3,5,6)$ or on OST $(j=2,4)$ is then given by

$$\delta_{j}^{inj}=1-F_{inj}(1-(V_{j}^{S}e_{s}+\left( 1-V_{j}^{S} \right)e_{u})$$

##### HIV disease progression - $P_{i,j,k}^{m,n}$

These terms are concerned with HIV disease progression and are denoted by $P_{i,j,k}^{m,n}$. Disease progression for those not on ART $(k=1)$ is given by

$$P_{i,j,k}^{2,n}=-\tau_{A}X_{i,j,k}^{2,n}$$

$$P_{i,j,k}^{3,n}=\tau_{A}X_{i,j,k}^{2,n}-{\tau_{E}^{k}\tau}_{C}X_{i,j,k}^{3,n}$$

$$P_{i,j,k}^{4,n}={\tau_{E}^{k}\tau}_{C}X_{i,j,k}^{3,n}-\tau_{E}^{k}\tau_{P}X_{i,j,k}^{4,n}$$

$$P_{i,j,k}^{5,n}={\tau_{E}^{k}\tau}_{P}X_{i,j,k}^{4,n}-\tau_{E}^{k}\tau_{I}X_{i,j,k}^{5,n}$$

where

- $\tau_{A}$ is the rate of progressing from acute HIV infection to latent HIV infection
- $\tau_{C}$ is the rate of progressing from latent HIV infection to pre-AIDS HIV infection if not on ART
- $\tau_{P}$ is the rate of progressing from pre-AIDS HIV infection to AIDS if not on ART
- $\tau_{I}$ is mortality due to AIDS if not on ART
- $\tau_{E}^{k}$ is the relative reduction in HIV disease progression and mortality due to AIDS if on ART (note that $\tau_{E}^{1}=1$).

##### Transitions on and off ART - $K_{i,j,k}^{m,n}$

We assume that those in the latent, pre-AIDS and AIDS phases of HIV infection are able to enrol on ART and are lost-to follow-up from ART.

$$K_{i,j,1}^{m,n}=-O_{j}\eta X_{i,j,1}^{m,n}+L_{j}\zeta X_{i,j,2}^{m,n} if m=3,4,5$$

$$K_{i,j,2}^{m,n}=O_{j}\eta X_{i,j,1}^{m,n}-L_{j}\zeta X_{i,j,2}^{m,n} if m=3,4,5$$

$$K_{i,j,k}^{m,n}=0 if m=1,2$$

where

- $\eta$ is the rate of enrolment onto ART if in the latent HIV, pre-AIDS and AIDS stages of infection.
- $\zeta$ is the rate of loss to follow up from ART if in the latent HIV, pre-AIDS and AIDS stages of infection
- $O_{j}$ is the effect of being on OST on ART enrolment (i.e. $O_{j}=1$ when $j=1,3,5,6$).
- $L_{j}$ is the effect of being on OST on loss-to follow-up of ART (i.e. $L_{j}=1$ when $j=1,3,5,6$).

##### HCV transmission and treatment - $\Pi_{i,j,k}^{m,n}$

These terms are concerned with HCV transmission and are denoted by $\Pi_{i,j,k}^{m,n}$

$$\Pi_{i,j,k}^{m,1}=-\beta_{j}X_{i,j,k}^{m,1} \forall m$$

$$\Pi_{i,j,k}^{1,2}=-\left( 1-\alpha_{-} \right)\beta_{j}X_{i,j,k}^{1,2}+\alpha_{-}{\beta_{j}X}_{i,j,k}^{1,1}+s\omega X_{i,j,k}^{1,4}$$

$$\Pi_{i,j,k}^{m,2}=-\left( 1-\alpha_{+} \right)\beta_{j}X_{i,j,k}^{m,2}+\alpha_{+}\beta_{j}X_{i,j,k}^{m,1}+s\omega X_{i,j,k}^{m,4} m\geq2$$

$$\Pi_{i,j,k}^{1,3}=\left( 1-\alpha_{-} \right)\beta_{j}\left( X_{i,j,k}^{1,1}+X_{i,j,k}^{1,2} \right)-\Psi X_{i,j,k}^{1,3}+\left( 1-s \right)\omega X_{i,j,k}^{1,4}$$

$$\Pi_{i,j,k}^{m,3}=\left( 1-\alpha_{+} \right)\beta_{j}\left( X_{i,j,k}^{m,1}+X_{i,j,k}^{m,2} \right)-\Psi X_{i,j,k}^{m,3}+\left( 1-s \right)\omega X_{i,j,k}^{m,4} m\geq2$$

$$\Pi_{i,j,k}^{m,4}=\Psi X_{i,j,k}^{m,3}-\omega X_{i,j,k}^{m,4} \forall m$$

where

- $\beta_{j}$ is the HCV force of infection for individuals accessing harm reduction state $j$.
- $\alpha_{-}$ is the proportion of HCV infections that spontaneously clear among HIV negative PWID.
- $\alpha_{+}$ is the proportion of HCV infections that spontaneously clear among HIV positive PWID.
- $\Psi$ is the HCV treatment rate
- $s$ is the sustained viral response rate among those undertaking HCV treatment
- $1/\omega$ is the average duration of HCV treatment.

HCV force of infection

The HCV force of infection for PWID in each intervention state $j$ is denoted by $\beta_{j}$ and is given by:

$$\beta_{j}={\Phi_{j}^{HCV}\beta}_{inj}^{HCV}\frac{C_{-}+C_{+}}{T}$$

where

$$C_{-}=\sum_{i} \sum_{j} \sum_{k} {\Phi_{j}^{HCV}X}_{i,j,k}^{1,3}$$

$$C_{+}=\sum_{i} \sum_{j} \sum_{k} \sum_{m\geq2} {F\Phi}_{j}^{HCV}X_{i,j,k}^{m,3}$$

$$T=\sum_{i} \sum_{j} \sum_{k} \sum_{m} \sum_{n} \Phi_{j}^{HCV}X_{i,j,k}^{m,n}$$

where

- $\beta_{inj}^{HCV}$ is the HCV transmission rate for PWID who are not currently accessing OST or NSP and are susceptible to HIV.
- $\Phi_{j}^{HCV}$ denotes the relative reduction in HCV transmission for injecting transmission if accessing OST ($j=2$), NSP $(j=3,6)$ or both $\left( j=4 \right)$. Note that when not accessing OST or NSP $(j=1,5)$ $\Phi_{j}^{HCV}=1$.
- $F$ is the relative increase in HCV transmissibility if PWID are HIV positive compared to HIV negative.

##### Non-HIV related mortality and injecting cessation - $M_{i,j,k}^{m,n}$

These terms are concerned with cessation of injecting and non-HIV related mortality.

$$M_{i,j,k}^{m,n}=-\left( \mu_{i,1}+\mu_{2} \right)X_{i,j,k}^{m,n}$$

where

- $\mu_{i,1}$ is the non-HIV related mortality rate for gender $i$.
- $\mu_{2}$ denotes the rate of cessation of injecting.

### Injecting Risk Behaviours among PWID

**Appendix Table 1:** Injecting risk behaviours amongst male and female PWID from the TLC-IDU study.

|  | **Nairobi** | | **Coastal region** | |
| --- | --- | --- | --- | --- |
|  | **Males**  **(95% CI)** | **Females**  **(95% CI)** | **Males**  **(95% CI)** | **Females**  **(95% CI)** |
| Average number of times injecting drugs in last 30 days | 66.7  (65.1-68.4) | 63.8  (60.9-66.7) | 65.1  (63.4-66.9) | 71.5  (66.5-76.5) |
| Average number of days injected drugs in last month. | 27.8  (27.3-28.3) | 27.1  (26.4-27.9) | 26.2  (25.8-26.6) | 27.3  (26.6-28.1) |
| Average number of times injected on a typical day when injecting drug use occurred. | 2.5  (2.4 – 2.6) | 2.3  (2.2-2.4) | 2.5  (2.4-2.5) | 2.7  (2.5-2.8) |
| Percentage that at the last injection used a syringe/needle that was previously used by someone else. | 10.3%  (8.5-12.2) | 14.5%  (9.9-19.1) | 2.4%  (2.0-2.9%) | 5.7%  (0.8-10.5%) |

### Model Parameterisation

**Appendix Table 2: Prior ranges for model parameters that differ between Nairobi and the Coastal region**

| **Parameter** | **Prior parameter range** | | **Source** |
| --- | --- | --- | --- |
|  | **Nairobi** | **Coastal region** |  |
| Average duration of injecting (years) | 1.75-7.0 | 2.05-8.2 | TLC-IDU[3] |
| HIV prevalence amongst new male PWID in 2012 | 3.4-6.4% | 3.1-5.4% | TLC-IDU[3] |
| HIV prevalence amongst male adults in 2012 | 2.4-5.2% | 1.1-4.0% | KAIS 2012[4] |
| HIV prevalence amongst new female PWID in 2012 | 12.2-26.1% | 27.9-50.1% | TLC-IDU[3] |
| HIV prevalence amongst female adults in 2012 | 4.2-8.0% | 4.1-8.1% | KAIS 2012[4] |
| Proportion of PWID on ART that are virally supressed | 28-40% | 35-48% | TLC-IDU[3] |
| Mean log HIV viral load if on ART and virally suppressed | 2.70-2.74 | 2.66-2.70 | TLC-IDU[3] |
| Mean log HIV viral load if on ART and not virally suppressed | 3.71-4.00 | 3.81-4.05 | TLC-IDU[3] |
| Mean log HIV viral load if not on ART and not virally suppressed | 4.22-4.42 | 4.09-4.32 | TLC-IDU[3] |
| Proportion of male PWID’s partners that are PWID | 0.11-0.17 | 0.12-0.19 | TLC-IDU[3] |
| Proportion of female PWID’s partners that are PWID | 0.34-0.50 | 0.24-0.38 | TLC-IDU[3] |

**Appendix Table 3: Prior ranges for parameters used in the model that do not differ between Nairobi and the Coastal region**

| Parameter | | Prior parameter distribution | Source |
| --- | --- | --- | --- |
| PWID related parameters | | | |
| Non-HIV mortality among PWID (/year) | | Normal: 3.53% (95%CI: 2.81 – 4.24) | [5] Used to calculate mortality amongst male and female PWID. |
| Ratio of crude mortality rates in male versus female PWID | | Normal: 1.32 (95%CI: 1.21-1.44) | [5] Used to calculate mortality amongst male and female PWID. |
| Year injecting drug use started $\boldsymbol{(}\boldsymbol{Yr}_{\boldsymbol{inj}}\boldsymbol{)}$ | | Uniform: 1997 - 2001 | [6] |
| Number of male PWID in $\boldsymbol{Yr}_{\boldsymbol{inj}}$ | | Uniform:  8000–20000 | Uninformative prior |
| Relative number of female PWID in $\boldsymbol{Yr}_{\boldsymbol{inj}}$ compared to male PWID | | Uniform:  0.05–0.4 | Range assumed based on prior knowledge of proportion of PWID population that are male. |
| HIV related parameters | | | |
| Average duration of the acute stage of HIV infection (months) | | Triangular: 2.9 (1.23-6.0) | [7] |
| Average duration of the pre-AIDs stage of HIV infection (months) | | Triangular: 9 (4.81-14) | [7] |
| Time to AIDs (Years) | | Triangular: 9.4 (5.5-10.1) | [8] Used to calculate average duration of the chronic HIV stage. |
| Time to death from AIDS (months) | | Lognormal: 10 (95%CI: 6.79 – 12.7) | [8] |
| Transmissibility in acute stage | | Lognormal: 276 (95%CI: 131-509) | [7] |
| Transmissibility in chronic stage | | Lognormal: 10.6 (95%CI: 7.61-13.3) | [7] |
| Transmissibility in Pre-Aids stage | | Lognormal: 76 (95% CI: 41.3-128.0) | [7] |
| Increase in HIV transmissibility per log increase in HIV viral load. | | Lognormal: 2.45 (95%CI 1.85-3.26) | [1] |
| HIV sexual and injecting transmission parameters | | | |
| HIV sexual transmission rate from males to females | | Uniform: 0–0.5 | Uninformative prior |
| HIV sexual transmission rate from females to males | | Uniform: 0–0.5 | Uninformative prior |
| HIV injecting transmission rate | | Uniform: 0–0.5 | Unknown, varied to calibrate model to HIV prevalence trends |
| ART related parameters | | | |
| Reduction in HIV progression if on ART | | Uniform: 0.20-0.30 | [9] |
| Relative ART coverage amongst individuals initiating injecting vs established PWID | | Uniform:  0.8–1.2 | Assumption |
| Annual increase in ART enrolment rate (/year) over | |  | Unknown, varied to calibrate the model to ART coverage in 2011 and 2015 and then stable after 2015 |
|  | 2004-June 2011 | Uniform: 0–1 |  |
|  | June 2011 – March 2015 | Uniform: 0–1 |  |
| ART LTFU amongst the general population | | Uniform: 0.04 - 0.37 | [10-15] |
| Relative risk of ART LTFU amongst PWID vs general population. | | Lognormal: 1.36 (95%CI: 1.22-1.52) | [16] |
| Relative effectiveness of ART on reducing transmission risk for injecting compared to reducing sexual transmission risk | | Uniform: 0.7–1 | Assumption as less evidence of the effectiveness of ART on injecting transmission than sexual transmission. |
| HCV related parameters | | | |
| Seeded HCV prevalence | | 0.2% |  |
| HCV injecting transmission rate | | Uniform: 0.0001–2 | Unknown, varied to calibrate to prevalence estimates |
| Increase in HCV transmissibility if HIV positive | | Uniform: 1 - 7 | [17] |
| Proportion of HCV infections amongst HIV negatives that spontaneously clear | | Uniform: 0.22 - 0.29 | [18] |
| Proportion of HCV infections amongst HIV positives that spontaneously clear | | Uniform: 0.115 - 0.193 | [19] |
| Proportion of HCV treatments that result in SVR | | 90.1% | [20] |
| OST/NSP related parameters | | | |
| OST exit rate (/year) | | Uniform: 0.1-2 | Uninformative prior – calibrated to number ever and currently on OST |
| Annual increase in NSP enrolment rate over 2013-2017.75 | | Uniform:0-1 | Unknown, varied to calibrate model to NSP trends over time |
| NSP exit rate | | Uniform: 0–2 | Unknown, varied to calibrate model to NSP trends over time |
| Relative reduction in the risk of HCV transmission if on OST | | Lognormal: 0.50 (95%CI: 0.40-0.63) | [21] |
| Relative reduction in the risk of HCV transmission if on NSP | | Lognormal: 0.44 (95%CI: 24-0.80) | [21] |
| Relative reduction in the risk of HIV transmission if on OST | | Lognormal: 0.46 (95%CI: 0.32-0.67) | [22] |
| Relative reduction in the risk of HIV transmission if on NSP | | Lognormal: 0.42 (95%CI: 0.22-0.81) | [23] |
| Increase in odds of being virally supressed when on ART if on OST vs off OST | | Lognormal: 1.45 (95%CI: 1.21-1.73) | [24] |
| Increase in ART initiation rate if on OST vs off OST | | Lognormal: 1.87 (95%CI: 1.50-2.33) | [24] |
| Decrease in ART LTFU if on OST vs off OST | | Lognormal: 0.77 (95%CI: 0.63-0.95) | [24] |

### Model Calibration

The model was calibrated using an approximate Bayesian computation Sequential Monte Carlo (ABC SMC) method[25] to data on the: (1) HIV prevalence amongst male and female PWID from each round of the TLC study; (2) HCV antibody prevalence amongst PWID from round 6 of the TLC study; (3) self-reported ART coverage amongst HIV positive PWID from each round of the TLC study; (4) programme data on the number of PWID currently or ever enrolled onto OST; (5) programme data on the number of PWID recently in contact with NSP; (6) PWID population size estimates; (7) estimates of proportion of PWID that are female from the TLC study and (8) bounds for the sexual HIV incidence among male and female PWID. Appendix table 4 shows the calibration data used in the SMC routine. During the ABC SMC, 5,000 parameter sets were sampled from prior probability distributions (Appendix – Tables 2-3). These were then perturbed and resampled in an iterative process to achieve 5,000 parameter sets where

- PWID population size was between the bounds (Table 1)
- The mean square difference of the number of people on OST and NSP between the model and data estimates were improved on by < 2% of the previous iteration.
- The likelihood of the model estimating the proportion of PWID that are female, HIV prevalence among males and females, and ART coverage were improved on by <2% of the previous iteration.
- HCV prevalence among all PWID was between the bounds (see Table 1)
- Sexual HIV incidence matched what would be expected based on prevalence when individuals start injecting.

We repeated the process of the ABC SMC for multiple sets of 5,000 parameter sets until the median of key model projections converged (were <5% of the previous combined sets). These key model projections were:

1. HIV prevalence among male PWID in 2019
2. HIV prevalence among female PWID in 2019
3. HCV prevalence among PWID in 2019
4. % of new HIV infections among male PWID in 2019 that is due to sexual transmission
5. % of new HIV infections among female PWID in 2019 that is due to sexual transmission
6. % of HIV infections averted by ART and harm reduction over 2004-2019 (i.e. since ART began)
7. % of HCV infections averted by harm reduction over 2013-2019 (i.e. since NSP began)

We then took a weighted sample (based on the likelihood of the run for HIV prevalence and ART coverage) to achieve a final parameter set, which was 10% of the full parameter runs. In Nairobi, 6 implementations of the ABC SMC was needed to reach convergence, producing 30,000 parameters sets giving 3,000 final parameter sets after sampling. In the Coastal region, 8 implementations of the ABC SMC was needed to reach convergence, producing 40,000 parameters sets giving 4,000 final parameter sets after sampling.

**Appendix Table 4**: Data used in the ABC SMC fitting method to calibrate the model for Nairobi and the Coastal region.

|  | **Nairobi** | **Coastal region** | **Date of Estimate** | **Data Source** |
| --- | --- | --- | --- | --- |
| **Proportion of PWID that are female** | 14.7% (95% CI: 13.1-16.4) | 10.6% (95% CI: 9.2-11.9%) | 2012-2015 | TLC-IDU[3] |
| **PWID population Size** | 9750-17150 | 5700-13100 | 2013 | [26] |
| **HIV prevalence amongst male PWID** | 15.4% (95%CI: 10.4–23.1)  10.2% (95% CI: 8.2-12.1)  14.9% (95% CI: 10.1-19.6)  16.6% (95% CI: 12.9-20.3)  12.1% (95% CI: 10.6-13.6)  12.6% (95% CI: 10.7-14.5)  9.6% (95% CI: 8.2-11.0) | N/A  16.5% (95% CI: 13.8-19.3)  14.9% (95% CI: 11.5-18.4)  14.7% (95% CI: 11.9-17.5)  13.9% (95% CI: 10.2-17.5)  11.9% (95% CI: 8.4-15.5)  13.9% (95% CI: 10.3-17.4) | Feb 2011  Sept 2012  July 2013  March 2014  Sept 2014  March 2015  Sept 2015 | 2011 IBBA[27]  TLC-IDU[3]  TLC-IDU[3]  TLC-IDU[3]  TLC-IDU[3]  TLC-IDU[3]  TLC-IDU[3] |
| **HIV prevalence amongst female PWID** | 60.7% (95%CI: 14.6–87.2)  34.2% (95% CI: 23.4-44.9)  37.9% (95% CI: 25.5-50.3)  36.1% (95% CI: 21.7-50.5)  45.1% (95% CI: 36.8-53.4)  29.5% (95% CI: 22.5-36.4)  29.1% (95% CI: 19.8-38.4) | N/A  52.2% (95% CI: 40.6-63.8)  43.9% (95% CI: 30.2-57.5)  43.5% (95% CI: 31.8-55.3)  45.2% (95% CI: 35.2-55.2)  43.3% (95% CI: 25.1-61.5)  48.1% (95% CI: 31.2-65.0) | Feb 2011  Sept 2012  July 2013  March 2014  Sept 2014  March 2015  Sept 2015 | 2011 IBBA[27]  TLC-IDU[3]  TLC-IDU[3]  TLC-IDU[3]  TLC-IDU[3]  TLC-IDU[3]  TLC-IDU[3] |
| **ART coverage amongst HIV positive PWID** | 16.5% (95% CI: 8.8-24.2)  30.6% (95% CI: 15.6-45.4)  33.2% (95% CI: 23.4-42.9)  41.1% (95% CI: 34.1-48.0)  47.9% (95% CI: 42.6-53.2)  65.7% (95% CI: 60.3-71.0) | 51.8% (95% CI: 43.0-60.7)  50.9% (95% CI: 38.9-63.0)  50.4% (95% CI: 40.7-60.1)  54.4% (95% CI: 41.0-67.8)  64.0% (95% CI: 49.9-78.1)  64.3% (95% CI: 50.1-78.4) | Sept 2012  July 2013  March 2014  Sept 2014  March 2015  Sept 2015 | TLC-IDU[3]  TLC-IDU[3]  TLC-IDU[3]  TLC-IDU[3]  TLC-IDU[3]  TLC-IDU[3] |
| **HCV antibody prevalence amongst PWID** | 10.9% (95% CI: 8.4-13.3)  N/A | 23.6% (95% CI: 19.5-27.7)  35.8% (95% CI: 31.0-40.7) | Sept 2015  2015 | TLC-IDU[3]  KANCO unpublished study |
| **Number of PWID on NSP** | 1267  2094  1730  2292  2986  3222  2909  3090  4151  3373  3617  5234  5111  4642  4597  5107  7799 | 1430  1887  1429  2680  1831  2276  2116  2342  2240  4168  4558  4568  3510  4536  3569  4584  4586 | Aug 2013  Nov 2013  Feb 2014  May 2014  Aug 2014  Nov 2014  Feb 2015  May 2015  Aug 2015  Nov 2015  Feb 2016  May 2016  Aug 2016  Nov 2016  Feb 2017  May 2017  Aug 2017 | Programme data |
| **Number of PWID ever enrolled onto OST** | 45  78  138  192  228  281  338  397  457  499  534  554  570  582  597  597  597  597  597  597  597  597  597  597  597  633  672  673  677  677  731  733  808  808  810  843 | 0  11  82  138  145  145  145  145  193  251  294  338  386  441  491  523  573  632  709  814  859  947  1014  1023  1060  1139  1177  1178  1178  1178  1229  1288  1427  1432  1506  1555 | Jan 2015  Feb 2015  March 2015  April 2015  May 2015  June 2015  July 2015  Aug 2015  Sept 2015  Oct 2015  Nov 2015  Dec 2015  Jan 2016  Feb 2016  March 2016  April 2016  May 2016  June 2016  July 2016  Aug 2016  Sept 2016  Oct 2016  Nov 2016  Dec 2016  Jan 2017  Feb 2017  March 2017  April 2017  May 2017  June 2017  July 2017  Aug 2017  Sept 2017  Oct 2017  Nov 2017  Dec 2017 | Programme data |
| **Number currently enrolled on OST** | 500 | N/A | 2018 | Programme data |
| **Average years of sexual risk before males start injecting** | 14.3-15.3 | 15.3-16.0 | 2013 | TLC-IDU[3] |
| **Average years of sexual risk before females start injecting** | 12.8-14.7 | 11.0-13.9 | 2013 | TLC-IDU[3] |
